# Impaired Probabilistic Learning deficits in Schizophrenia: A study with Motor Execution and Imagery

**DOI:** 10.64898/2026.07.03.26357176

**Authors:** Yanina Leon Uscapi, Patricia Silva de Camargo, Paulo Roberto Cabral-Passos, Renan Biokino, July Silveira Gomes, André Frazão Helene, Ary Gadelha, Dulce Barbosa

**Affiliations:** Paulista School of Nursing, Federal University of São Paulo, São Paulo, Brazil; Institute of Biosciences, University of São Paulo, São Paulo, Brazil; Institute of Mathematics and Statistics, University of São Paulo, São Paulo 05508-090, Brazil; Department of Psychiatry, Federal University of São Paulo, São Paulo, Brazil

**Keywords:** schizophrenia, implicit learning, probabilistic motor learning, motor imagery, motor execution, serial reaction time task, cognitive deficits

## Abstract

Schizophrenia is associated with cognitive impairments, including deficits in implicit learning. Probabilistic serial reaction time tasks (SRTT) offer an objective approach to characterizing these deficits through both motor execution (ME) and motor imagery (MI), the mental simulation of movement without physical action. Whether implicit probabilistic sequence learning is impaired across both modalities in schizophrenia remains poorly understood. Thirty individuals with schizophrenia (ME: n=12; MI: n=13) and 40 healthy controls (ME: n=20; MI: n=20) completed an auditory probabilistic SRTT. Symptom severity was assessed with the PANSS and cognitive functioning with the MCCB. Healthy controls demonstrated a robust signature of implicit probabilistic sequence learning, whereas participants with schizophrenia exhibited weaker and less consistent learning signatures, particularly during motor imagery. Sensitivity to probabilistic structure differed significantly between groups during motor execution but not motor imagery. Participants with schizophrenia also showed significantly longer reaction times than controls across both modalities, consistent with generalized psychomotor slowing. Greater PANSS-General severity was associated with greater deviation from the probabilistic learning patterns observed in healthy controls during ME, whereas higher MCCB verbal learning scores were associated with greater similarity to these learning patterns during MI. These findings indicate that implicit probabilistic sequence learning is impaired in schizophrenia across both motor execution and motor imagery, and that these deficits are meaningfully associated with clinical symptom severity and cognitive functioning.

## 1. Introduction

Schizophrenia is a chronic and severely disabling psychiatric disorder that consistently affects approximately 23 million people worldwide (Vos et al., 2017; Gadelha et al., 2025; WHO, 2026). Its clinical presentation encompasses positive, negative, and cognitive symptoms, with cognitive impairments representing a major determinant of functional outcome (WHO, 2026). Among these cognitive impairments, alterations in implicit learning and memory have been increasingly recognized as early and pervasive features of the disorder.

Implicit learning, defined as the acquisition of knowledge without conscious awareness of what has been learned, can be assessed through non-verbal paradigms (Spataro et al., 2016). One widely used approach is the Serial Reaction Time Task (SRTT), in which participants respond to sequentially presented stimuli according to a predefined stimulus–response mapping (Kalra et al., 2019; Keele et al., 2003). Although the underlying sequence structure is typically not disclosed, participants progressively improve their performance by extracting regularities embedded in the stimulus stream. These regularities may range from deterministic to probabilistic, with the latter placing greater demands on the integration of statistical contingencies over time (Oliveira et al., 2024). The ability to encode transition probabilities, even in seemingly random sequences, is commonly referred to as statistical learning and relies on implicit memory processes (Hasher and Zacks, 1984).

SRTTs have long been used to assess implicit learning in individuals with schizophrenia (e.g., Siegert, Weatherall, and Bell, 2008; Chrobak et al., 2023). Siegert et al.’s (2008) meta-analysis addressed inconsistent findings across studies, concluding that individuals with schizophrenia exhibit distinct response patterns in SRTTs compared to healthy controls, specifically, a different relative order of reaction times across stimuli. These patterns have been linked to moderate impairments in implicit learning. However, Chrobak et al. (2023) recently argued that such distinct patterns, often interpreted as poorer performance, may instead reflect atypical temporal dynamics that warrant further investigation. Notably, these findings were consistent across task variants, indicating robustness to methodological differences in SRTT paradigms.

The temporal dynamics of SRTTs performance arise from the interplay between cognitive and sensory demands involved in detecting stimulus regularities and those required for coordinated motor execution. Isolating these independent contributions is critical for understanding the substrates underlying such dynamics. Motor imagery (MI), the mental simulation of movement in the absence of physical execution, offers a unique approach to this isolation. MI engages cortical circuits that closely resembles those recruited during overt execution (Decety et al., 1993; Oishi et al., 1994; Pascual-Leone et al., 1995; Crammond, 1997; Helene and Xavier, 2006), producing similar temporal dynamics (Jeannerod and Decety, 1995), while also exhibiting its own distinct temporal signature (de Camargo, Cabral-Passos, and Helene, 2026). Although motor imagery capacity has been tested in individuals with schizophrenia (Mazhari et al., 2015), the temporal dynamics of MI in this population lack investigation.

Based on these considerations, the present study aims to compare the temporal dynamics of SRTT performance between individuals with schizophrenia and healthy controls across both motor execution (ME) and motor imagery (MI) modalities. We hypothesized that individuals with schizophrenia would exhibit generalized psychomotor slowing in both ME and MI, alongside reduced sensitivity to probabilistic regularities, particularly in MI, where internally generated processes are more heavily engaged. This approach may help elucidate the cognitive substrates underlying performance differences between these populations. By directly comparing ME and MI, we further aim to disentangle the contributions of overt motor processes from those of covert simulation, providing insights into whether impairments in schizophrenia are modality-specific or reflect broader deficits in implicit learning mechanisms. Here, we present evidence that individuals with schizophrenia exhibit generalized psychomotor slowing under both ME and MI conditions, with distinct patterns of sensitivity to sequence structure across modalities.

## 2. Materials and Methods

### 2.1 Participants

The study was conducted at the Cognition Science Laboratory, Institute of Biosciences, University of São Paulo (IB-USP), and at the Center for Integrated Mental Health Care (CAISM), Brazil, a reference institution for psychiatric care, research, and education. All procedures were carried out in accordance with the Declaration of Helsinki and approved by the local ethics committees (CAAE 66812022000005505, Federal University of São Paulo - UNIFESP; CAAE 15274718.8.0000.5464, Institute of Biosciences at the University of São Paulo - IB-USP). Written informed consent was obtained from all participants prior to their inclusion in the study.

The schizophrenia sample comprised 30 individuals, randomly assigned to one of two modality groups: Motor Execution (ME; n = 16; 5 women; mean age = 29.06±9.20 years) and Motor Imagery (MI; n = 14; 2 women; mean age = 30.71± 6.48 years). Due to technical issues during data acquisition, including incomplete trial recordings for some participants, 5 individuals were excluded from the final analysis to ensure dataset integrity and consistency. The final schizophrenia sample comprised 12 participants in the ME modality group and 13 participants in the MI modality group. Detailed demographic characteristics of all participants in the Schizophrenia Group (SG) are provided in Table 1.

**Table 1.** Demographic characteristics of participants in the Schizophrenia Group and questionnaire information of the participants separated into modality. Mean and standard deviations (SD) are given.

| | Motor Execution<br>(Mean ± SD ) | Motor Imagery<br>(Mean ± SD) | Total<br>(Mean ± SD) |
| --- | --- | --- | --- |
| Age (years) | 29.06±9.20 | 30.71±6.48 | 29.83±7.82 |
| Gender (F/M) | F(5); M(11) | F(2); M(12) | F(7); M (23) |
| Years Since Diagnosis | 8.81±7.81 | 10.78±7.52 | 9.73± 7.48 |
| Education (years) | 13.31±4.12 | 12.92±3.17 | 13.13±3.65 |
| Employment Status (U/E) | U(12); E(4) | U(11); E(3) | U(23); E(7) |
| Edinburgh Inventory | 77.29±13.88 | 80.95±28.71 | 79.12± 21.74 |
| KVIQP-10V | 30.93±7.92 | 30.52±7.92 | 30.73±7.72 |
| KVIQP-10C | 30.75±10.05 | 29.21±9.61 | 30.03±9.71 |

F = female; M = male; U = unemployed; E = employed; KVIQP-10V = Visual Motor Imagery Questionnaire (Brazilian version); KVIQP-10C = Kinesthetic Motor Imagery Questionnaire (Brazilian version).

Participants were eligible for the study if they met the following criteria: (1) age ≥ 18 years old; (2) had a diagnosis of schizophrenia according to DSM-5 (APA, 2013), confirmed via Axis I module (disorders) of the SCID-5 RV (2016); (3) according to their psychiatrist, received treatment for psychosis while being clinically stable; (4) right-handedness; (5) demonstrated ability to perform motor imagery tasks. Handedness was assessed using the *Edinburgh Handedness Inventory* (Oldfield, 1971), with scores > 50 indicating right-hand dominance. Motor imagery ability was evaluated using the *Kinesthetic and Visual Motor Imagery Questionnaire - KVIQP-10, Brazilian version* (Demanboro et al., 2018), which yields scores ranging from 9 to 45 per subscale, with higher scores indicating greater motor imagery ability. Participants scoring ≥ 25 were included in the study. Exclusion criteria comprised diagnosis of bipolar disorder (type I or II), Alzheimer’s disease, psychotic disorders other than schizophrenia, schizoaffective disorder, neurosyphilis, history of traumatic brain injury or stroke, substance use disorders, or hypothyroidism.

The control sample comprised 40 healthy individuals, also randomly assigned to two modality groups: Motor Execution (ME; n = 20; 12 women; mean age = 25.45 ±4.81 years) and Motor Imagery (MI = 20; 12 women; mean age = 26±4.74 years). Detailed demographic characteristics are presented in Table 2.

**Table 2.**
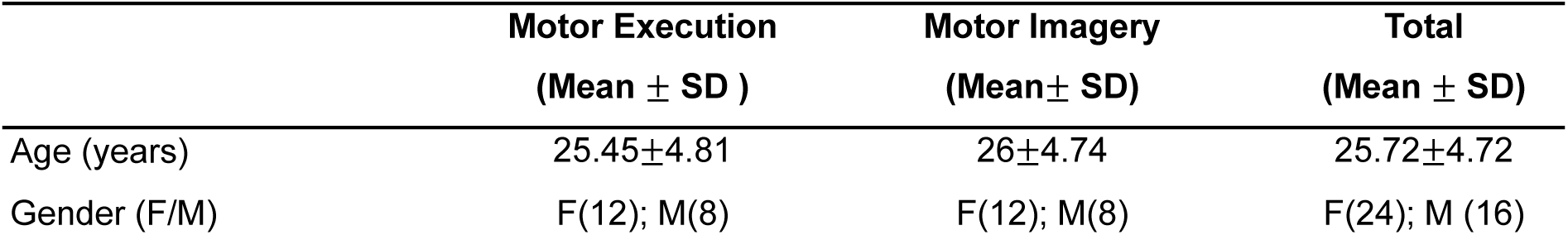
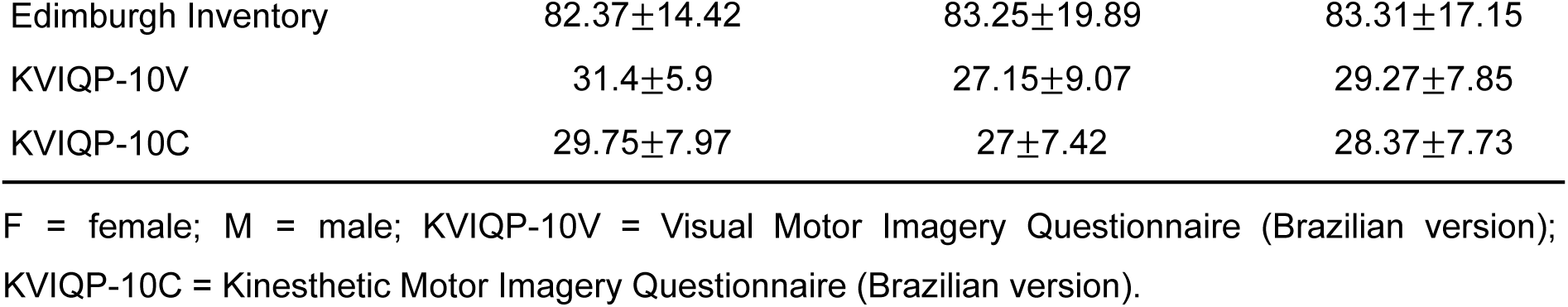
Demographic characteristics of participants in the Control Group and questionnaire information of the participants separated into subgroups. Mean and standard deviations (SD) are given.

| | Motor Execution<br>(Mean ± SD ) | Motor Imagery<br>(Mean ± SD) | Total<br>(Mean ± SD) |
| --- | --- | --- | --- |
| Age (years) | 25.45±4.81 | 26±4.74 | 25.72±4.72 |
| Gender (F/M) | F(12); M(8) | F(12); M(8) | F(24); M (16) |
| Edinburgh Inventory | 82.37±14.42 | 83.25±19.89 | 83.31±17.15 |
| KVIQP-10V | 31.4±5.9 | 27.15±9.07 | 29.27±7.85 |
| KVIQP-10C | 29.75±7.97 | 27±7.42 | 28.37±7.73 |

F = female; M = male; KVIQP-10V = Visual Motor Imagery Questionnaire (Brazilian version); KVIQP-10C = Kinesthetic Motor Imagery Questionnaire (Brazilian version).

Control participants were selected based on the following criteria: (1) age ≥ 18 years; (2) right-handedness as assessed by the Edinburgh Handedness Inventory; (3) ability to perform imaginative tasks as evaluated by the KVIQP-10; and (4) absence of any reported neurological, spinal, orthopedic, vascular, or muscular dysfunctions affecting the upper limb.

Symptom severity in participants with schizophrenia was assessed using the Positive and Negative Syndrome Scale (PANSS) (Kay, Fiszbein, & Opler, 1987; Higuchi et al., 2014), a 30-item, clinician-administered scale that evaluates three domains: Positive Symptoms (PANSS-P, 7 items), Negative Symptoms (PANSS-N, 7 items), and General Psychopathology (PANSS-G, 16 items). Each item is rated on a 7-point Likert scale (1 = absent to 7 = extreme), with higher scores indicating greater symptom severity. The Total PANSS score (PANSS-T) is computed as the sum of all 30 items. The PANSS is widely validated for its reliability and sensitivity in capturing the multidimensional nature of schizophrenia symptomatology, including hallucinations, delusions, blunted affect, and cognitive dysfunction. In the present study, PANSS subscales were used to explore potential associations between symptom severity and implicit learning performance. A summary of the participants’ scores is presented in Table S1.

The types and dosages of antipsychotic and other psychotropic medications used by participants in the Schizophrenia Group are detailed in Table S2. All participants were required to be on a stable medication regimen for at least four weeks prior to participation, in order to minimize potential acute effects of medication changes on cognitive performance. Medication adherence was monitored through self-report and clinical records. The study did not control for specific antipsychotic medications or dosages, as the primary focus was on the behavioral and cognitive outcomes of the experimental tasks.

Cognitive functioning in participants with schizophrenia was additionally assessed using the MATRICS Consensus Cognitive Battery (MCCB) (Nuechterlein et al., 2008), a standardized and comprehensive instrument designed to evaluate seven key cognitive domains: speed of processing, attention/vigilance, working memory, verbal learning, visual learning, reasoning/problem solving, and social cognition. In the present study, the social cognition domain was not administered given the low reliability coefficients reported for the Brazilian version of this domain (Fonseca et al., 2017). MCCB data is provided in Table S3.

### 2.2 Experimental Protocol

The experiment protocol and the probabilistic sequence used in the serial reaction time task (SRTT) were adapted from a previous study conducted by our research group (de Camargo, Passos-Cabral, and Helene, 2026). Auditory stimuli were presented using *Psychtoolbox-3 (Psychophysics Toolbox Version 3—PTB-3, CA, USA)* software version 3.0.18 Beta. All stimuli were recorded by a single native Brazilian Portuguese speaker to ensure consistency, captured using audio recording software (Audacity, version 3.0.0) with a USB conventional microphone (Blue Yeti, Logitech, Palo Alto, CA, USA), and were presented using Samsung in-ear headphones (Samsung model EO-EG920B, China). Data collection was conducted in a sound-attenuated, dimly lit room. Participants were seated approximately 50 cm from a 23-inch monitor (30 cm height × 52 cm width), and a keyboard was positioned comfortably in front of them. Standardized instructions were provided prior to the task, and all questions were addressed before testing began.

The task consisted of an auditory SRTT comprising three verbal stimuli, “one”, “two”, and “three” (in Brazilian Portuguese, *um*, *dois* and *três*), each corresponding to a specific motor response. On each trial, participants were required to respond as quickly as possible to the auditory stimulus by pressing a corresponding key with a specific finger. The stimulus-response mapping was as follows: “one” (left arrow key, index finger), “two” (down arrow key, middle finger), and “three” (right arrow, ring finger).

Participants in the ME group performed the task by physically pressing the keys as instructed. In contrast, participants in the MI group were instructed to mentally simulate the required response without making any movement, and upon completing the imagined action, they pressed the spacebar with their left thumb to indicate the end of their mental simulation (see de Camargo, Passos-Cabral, and Helene, 2026).

The stimulus sequence followed a probabilistic structure based on a context tree model, designed to assess implicit statistical learning. Each trial corresponded to one of three possible stimuli, with transitions determined by the probabilistic sequence structure. Transitions from stimulus “1” were always deterministic, with “1” always followed by “2”. For transitions from stimulus “2”, the outcome depended on the preceding stimulus: if “2” was preceded by “1”, the transition was probabilistic - either “3” (74% probability) or “2” (26% probability); if “2” was preceded by another “2”, the transition was deterministic, always followed by “1”. So one of the possible realizations of the stimuli sequence is “… 1 2 3 1 2 2 1 2 3 …”. For clarity, deterministic transitions were labeled as fixed (F; F1 and F2), and probabilistic transitions as variables (V; V2 and V3). A detailed description of the sequence generation is provided in de Camargo, Passos-Cabral, and Helene, (2026).

### 2.2 Statistical Analysis

Statistical analyses were conducted in Python using the Statsmodels (Seabold & Perktold, 2010) and Pingouin (Vallat, 2018) packages. Because reaction times were skewed, median values were used as the primary measure of central tendency: hereafter, ‘reaction times’ refers to both individual and median values. Normality was assessed using Shapiro–Wilk tests. When normality was violated in between-subjects comparisons, the Mann-Whitney U test was used. For analyses with two within-subjects variables, two-way repeated-measures ANOVA was applied; for analyses with one within and one between-subjects variable, a two-way mixed ANOVA was used. In relevant cases, variables were collapsed to enhance statistical power and interpretability. When normality violations persisted and no non-parametric test was available, rank transformations were applied to approximate normality. Sphericity violations were corrected using the Greenhouse–Geisser method. All analysis code, output, and detailed descriptions are publicly available at: https://github.com/PauloCabral-hub/Behavioral-Statistical-Learning-in-Schizophreny/tree/main/notebooks. Further methodological details are provided in the Supplementary Material.

## 3. Results

### 3.1 Comparing the magnitude of the response times in both groups

Figure 1 shows the distribution of reaction times for each group (Control and Schizophrenia), modality (Motor Execution - ME and Motor Imagery - MI), block (1–5), and stimulus type (F1, F2, V2, and V3). As illustrated, participants with schizophrenia exhibited higher overall mean reaction times compared to the control group. To formally assess this difference, we compared the distribution of individual reaction times for both groups, Control and Schizophrenia, using the Mann–Whitney U test, given that both distributions failed the normality assumption (p<0.01). The results supported the hypothesis of longer reaction times in Schizophrenia (n_1_ = 40, n_2_ = 25, U = 228,679.0, p < 0.001).

**Figure 1.**
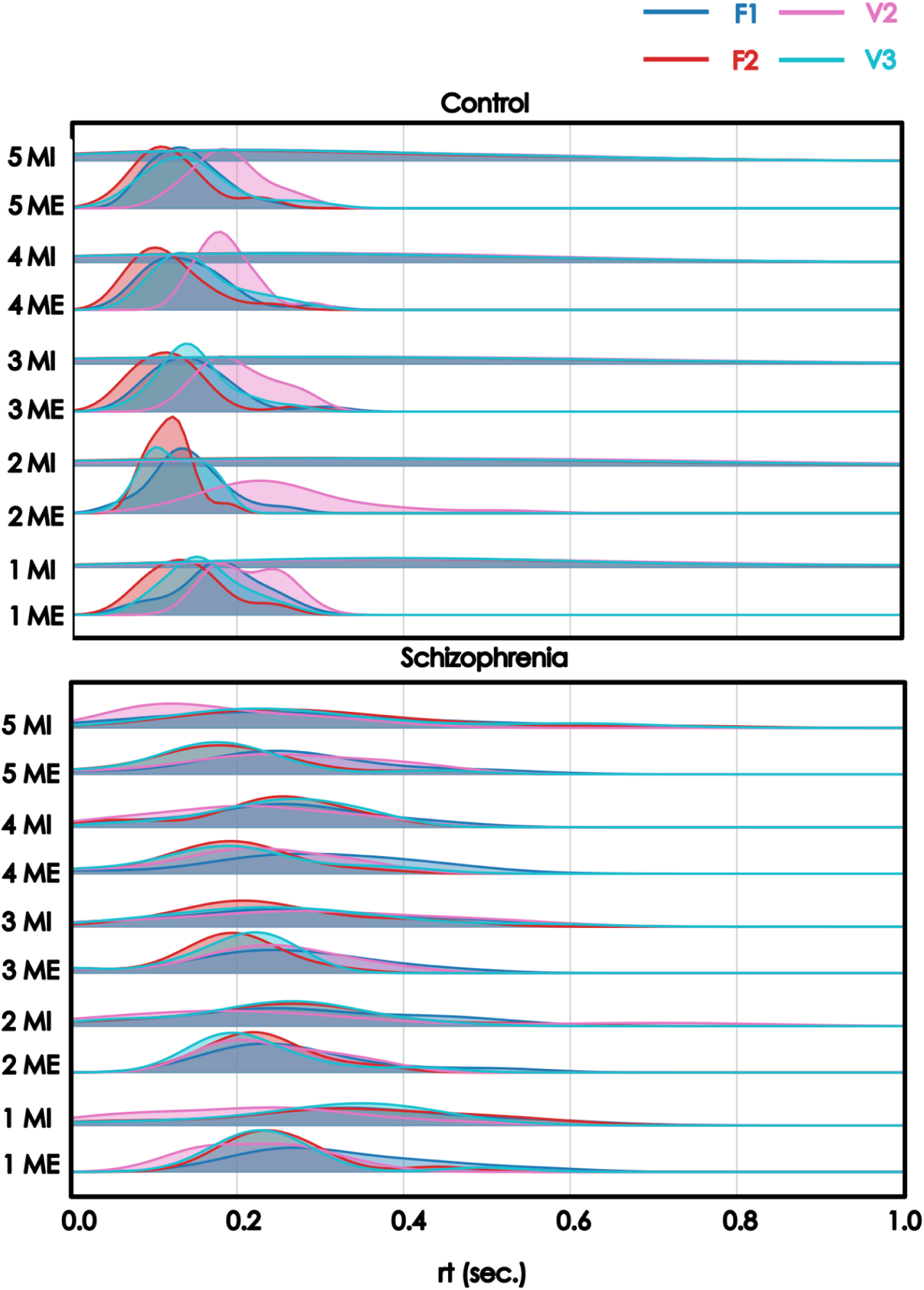
Generalized Psychomotor Slowing in Schizophrenia: Reaction Time Distributions by Modality, Block, and Stimulus Type. Response times for F1, F2, V2, and V3 are depicted in dark blue, red, magenta, and cian, respectively. Blocks are indicated on the left side of each graph (1-5), while the R and I indicate the motor execution and motor imagery, respectively.

### 3.2 Reaction times across blocks in Schizophrenia and Control groups

Figure 2 shows the distribution of the median reaction times (rt) for the Control group (top graphs) and Schizophrenia group (bottom graphs). Each distribution corresponds to a unique combination of levels from the variables Block and Stimulus Type. Each sample in the distribution represents the median reaction times (rt) of a single subject for that specific combination. A permutation-based repeated-measures ANOVA was used to evaluate the effects of Block, Stimulus Type, and their interaction on RTs. This approach was chosen because the normality assumption was violated across multiple distributions (see Supplementary Tables S4–S6)

**Figure 2.**
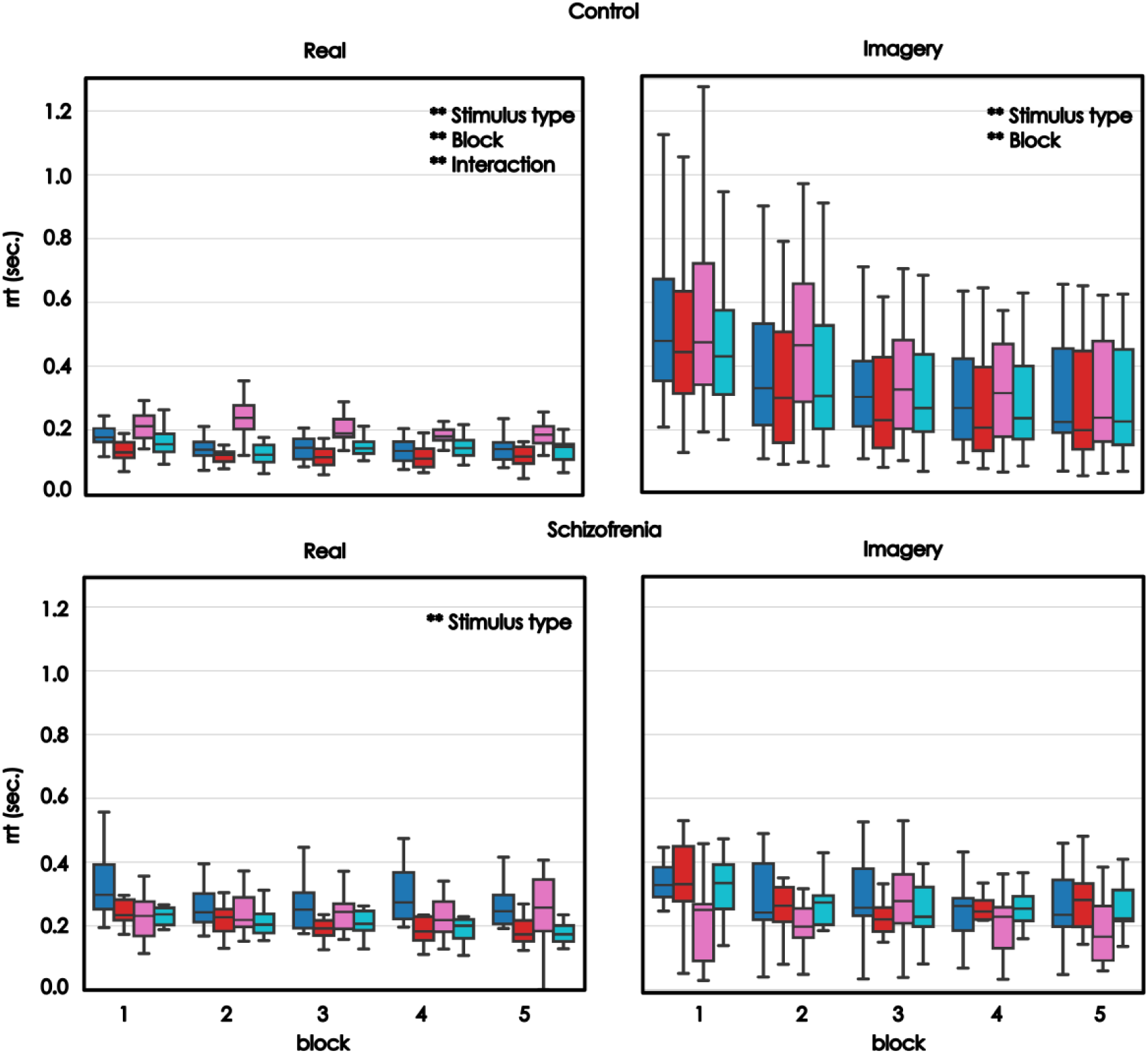
Distribution of the median reaction times (rt) by group, block, modality, and stimulus. Response times for F1, F2, V2, and V3 are depicted from left to right for each block (F1, F2, V2, and V3 are represented in dark blue, red, magenta, and cian, respectively). Global significant effects are shown on the top right corner of the graphs, * and ** indicates p < 0.01 in the repeated measures ANOVA permutation test.

For the Control participants under ME, a significant effect of stimulus type (F_(3,57)_ = 50.29, corrected *p* < 0.01), block (F_(4,76)_ = 4.24, corrected *p* < 0.01), and interaction between block and stimulus type (F_(12,228)_ = 7.05, corrected *p* < 0.01) were verified. For the Control group under MI, a significant effect of stimulus type (F_(3,57)_ = 6.13, corrected *p* < 0.01) and block (F_(4,76)_ = 5.32, corrected *p* < 0.01), while no significant interaction was found. For individuals with Schizophrenia under Execution modality, a significant effect was found only for stimulus type (F_(3,33)_ = 11.77, corrected *p* < 0.01). Under MI modality, no significant effects were detected.

### 3.3 Overall comparison of reaction times between Schizophrenia and Control groups

Figure 3 shows the distribution of median reaction times for each Stimulus Type and Modality when collapsing the variable Block. The graph on the left shows reaction times for the Control group, while the one on the right for the Schizophrenia group. A mixed-model ANOVA was applied to compare ME and MI within each group, taking stimulus type and modality as within-subjects and between-subjects factors, respectively. Each of the distributions corresponds to a unique combination of levels from variable Modality and variable Stimulus Type, with each sample in the distribution representing the median rt of a single subject for that specific combination. Within each graph, ME and MI are presented on the left and right, respectively. A rank-transformation procedure was adopted to mitigate violations of normality (details in the supplementary material, Table S7), assuming a small additional error as a consequence of persistent violations.

**Figure 3.**
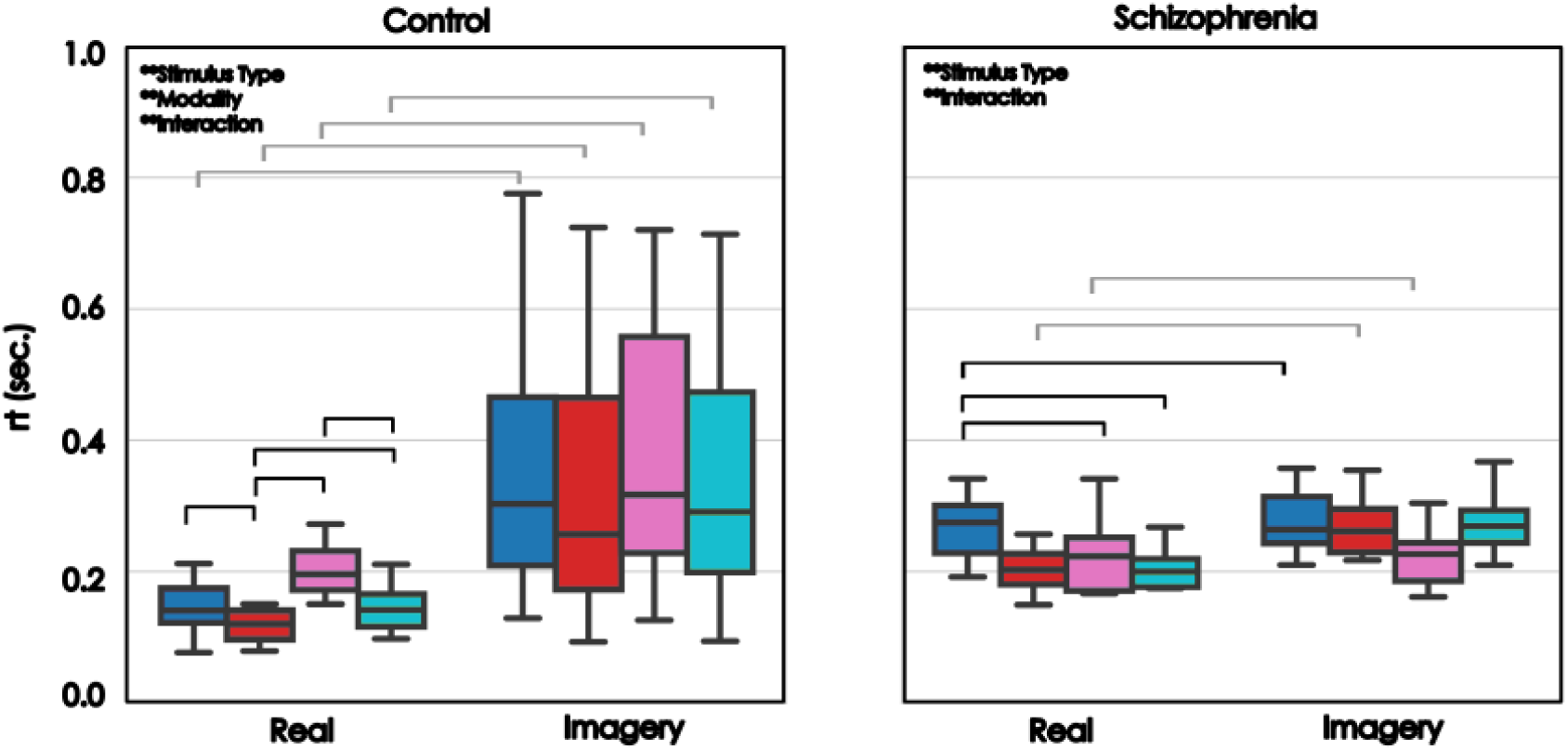
Distribution of the reaction times (rt) by modality and stimulus type. On the left for the Control group and on the right for the Schizophrenia group. Response times for F1, F2, V2 and V3 are depicted from left to right for each modality (F1, F2, V2, and V3 are represented in dark blue, red, magenta, and cian, respectively). Global significant effects are shown on the top left corner of the graphs, ** indicates p < 0.01 in the mixed ANOVA. Post-hoc pairwise comparisons are shown with black brackets (within subjects) and gray brackets (between subjects).

For the Control group, there were significant effects of the Stimulus Type (F_3,114_ = 56.029, p < 0.001), Modality (F_1,38_ = 27.05, p < 0.001), and the interaction between Stimulus Type and Modality (F_3,114_ = 18.49, p < 0.001). For the Schizophrenia group, significant effects were found only for Stimulus Type (F_3,69_ = 8.821, p < 0.001) and the interaction of Stimulus Type and Modality (F_3,69_ = 5.96, p < 0.001). Global effects are shown in Figure 3, top left corner, while significant post-hoc pairwise comparisons are shown with black brackets (within subjects) and gray brackets (between subjects).

Another mixed model ANOVA was used, this time, to compare both Control and Schizophrenia groups within each task modality. To illustrate the procedure, Figure 4 shows the same data as Figure 3, but now distributions are grouped within each graph by modality.

**Figure 4.**
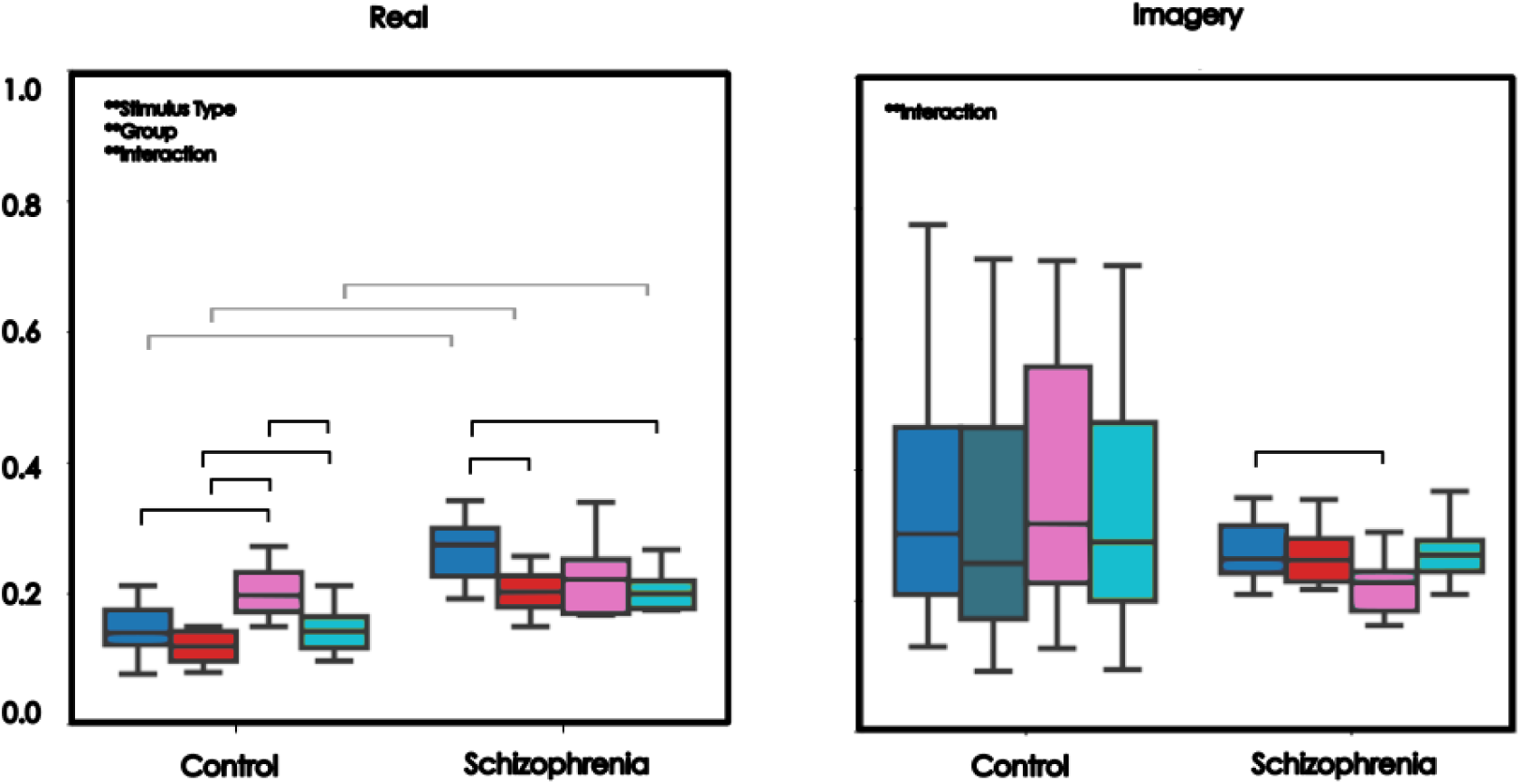
Distribution of the reaction times (rt) by group and stimulus type. On the left for ME (Real) and on the right for MI (Imagery). Response times for F1, F2, V2 and V3 are depicted from left to right for each modality (F1, F2, V2, and V3 are represented in dark blue, red, magenta, and cian, respectively). Global significant effects are shown on the top left corner of the graphs, ** indicates p < 0.01 in the mixed ANOVA. Post-hoc pairwise comparisons are shown with black brackets (within subjects) and gray brackets (between subjects).

In ME, significant effects were observed for Group (F_1,30_ = 16.23, p < 0.001), Stimulus Type (F_3,90_ = 34.08, p < 0.001), and their interaction (F_3,90_ = 20.44, p < 0.001). In MI, only the interaction between Stimulus Type and Group was significant (F_3,93_ = 11.61, p < 0.001). Global effects are shown in Figure 4 with post-hoc pairwise comparisons indicated with black brackets for within-subjects and gray brackets for between-subjects comparisons.

### 3.4 Relationship between reaction time patterns, clinical symptom severity, and cognitive functioning

Previous analyses revealed a marked difference between reaction times for F2 and V2 stimuli, particularly in the Control group under motor execution (Figures 3 and 4). Because this contrast captures one of the most prominent behavioral effects observed in the task, it was used as a summary metric for subsequent analyses. For each participant, blocks were collapsed and the percentage difference between the median reaction times of F2 and V2 was calculated as: 100 × (median reaction time of F2 / median reaction time of V2). Linear regression analyses were then performed between this metric and the Positive Symptoms (PANSS-P), Negative Symptoms (PANSS-N), General Psychopathology (PANSS-G), and Total PANSS (PANSS-T) scores.

Results are presented in Figures 5 for participants with schizophrenia under ME (blue) and MI (black) modalities, respectively. The graph includes the statistical description and the slope of the linear regression. Within the ME group, the relationship between PANSS subscales and the proposed metric for ME aligns with expectations. Specifically, as PANSS-P scores increase, the difference between F2 and V2 reaction times grows, becoming more similar to the pattern observed in the control group, though this relationship does not reach statistical significance. In contrast, both PANSS-N and PANSS-G exhibit an inverse linear relationship with the metric; that is, as negative symptoms and general psychopathology worsen, F2 and V2 reaction times become more similar (or even inverting their relative position), deviating from the pattern observed in the control group. Importantly, among these two subscales, only the PANSS-G reached nominal statistical significance (p < 0.05), although this did not remain significant after Benjamini–Hochberg correction for multiple comparisons. Notably, it also exhibited the steepest regression slope, suggesting a potentially meaningful association that may be confirmed in a larger sample. PANSS-T followed a similar trend but failed to reach significance. Under the MI modality, the same metric did not reveal any relevant relationship with PANSS subscales.

**Figure 5.**
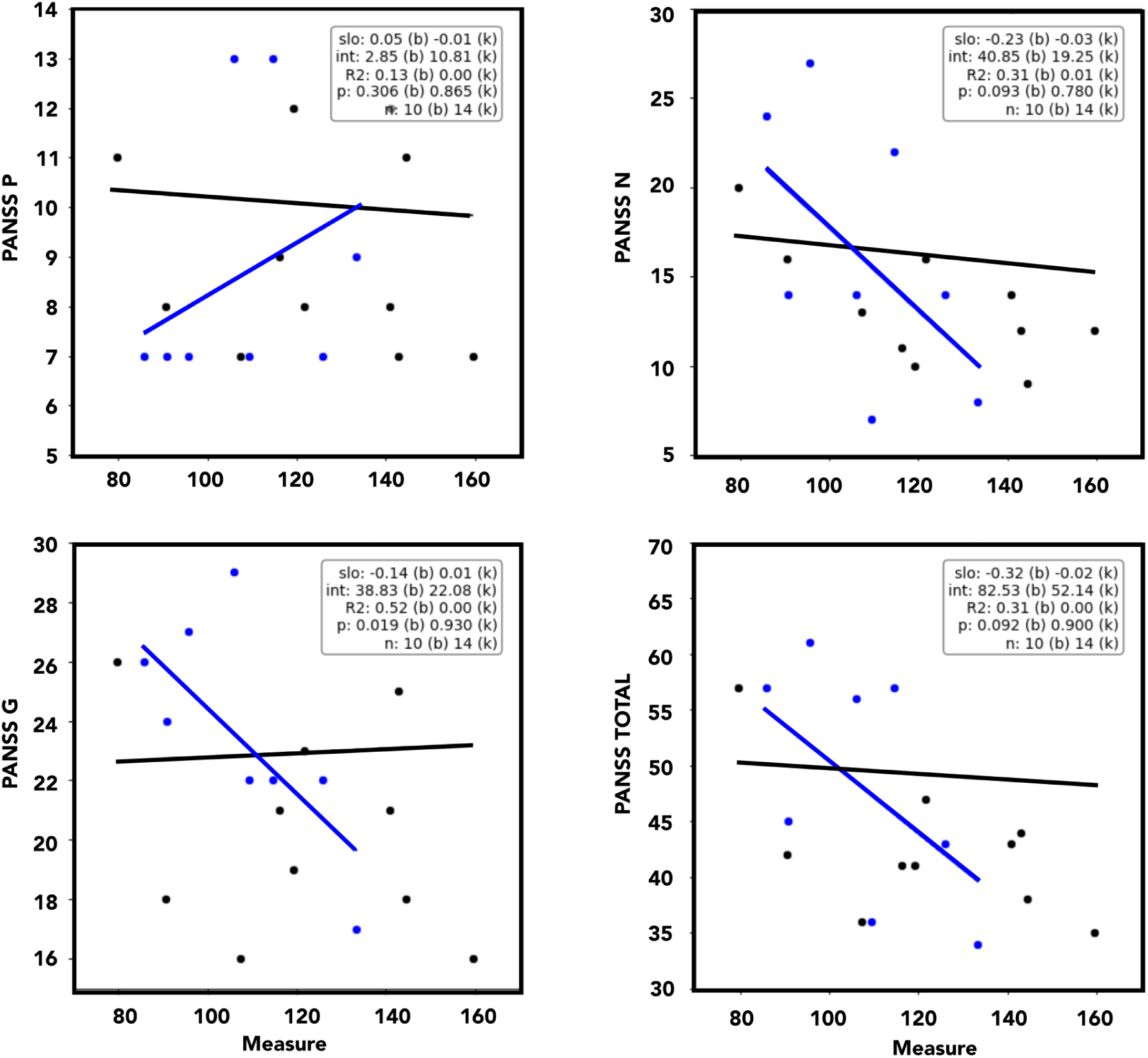
Linear regression analysis between PANSS (Positive and Negative Syndrome Scale) scores and the percentage change in median reaction time between F2 and V2 (indicated as measure) in the Motor Execution (ME, blue) and Motor Imagery (MI, black) modality groups of Schizophrenia patients. The dots represent individual data points associated with the corresponding linear regression. Significant correlations are indicated within the label on the top right corner (p < 0.05, b - ME, k - MI).

The same metric was applied in regression analyses against scores from the MATRICS Consensus Cognitive Battery (MCCB). The MCCB is commonly divided into the following cognitive domains: speed of processing, attention/vigilance, working memory, verbal learning, visual learning, reasoning/problem solving, and social cognition. For each of the six domains evaluated, a linear regression analysis was conducted to examine the relationship between our metric and the domain-specific MCCB scores. The results are presented in Figure 6. Notably, only the Motor Imagery (MI) condition showed a nominally significant correlation, specifically with the verbal learning domain (p < 0.05). However, this association did not remain significant after Benjamini–Hochberg correction for multiple comparisons and should therefore be interpreted with caution. This finding suggests that higher verbal learning scores are associated with higher values of our metric, corresponding more closely to the pattern observed in the control group during Motor Execution (ME).

**Figure 6.**
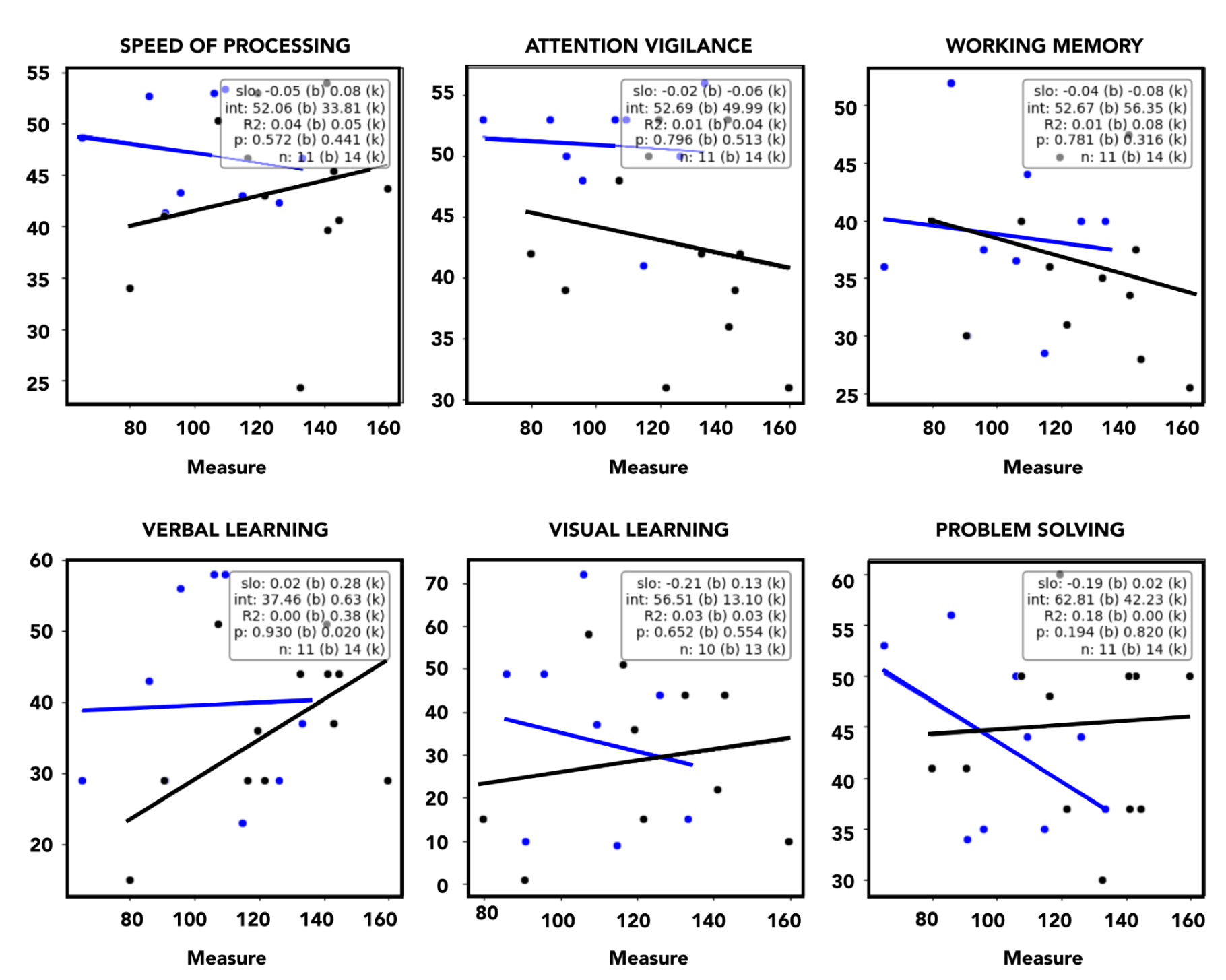
Linear regression analysis between MATRICS Consensus Cognitive Battery (MCCB) domain scores and the percentage change in median reaction time between F2 and V2 (indicated as measure) in the Motor Execution (ME, blue) and Motor Imagery (MI, black) modality groups of Schizophrenia patients. The dots represent individual data points associated with the corresponding linear regression. Significant correlations are indicated within the label on the top right corner (p < 0.05, b - ME, k - MI).

## Discussion

Overall, the results revealed clear differences between the Control and Schizophrenia groups. Healthy participants demonstrated evidence of implicit learning, characterized by progressive reductions in reaction times, sensitivity to probabilistic structure, and consistent modulation by stimulus type, particularly during motor execution. Although these effects were attenuated under motor imagery compared to motor execution, the findings suggest that internal simulation processes retain sensitivity to sequence-related regularities. In contrast, individuals with schizophrenia exhibited generalized response slowing (Fig. 1) and reduced sensitivity to probabilistic regularities. While some modulation by stimulus type was preserved during motor execution, this effect did not improve across practice blocks and the differences between stimulus types remained less pronounced, indicating limited adaptation over time. Under motor imagery, no consistent evidence of learning or stable stimulus-related modulation was observed. Together, these findings suggest that impairments in implicit probabilistic learning in schizophrenia are not restricted to motor execution processes, but may also involve difficulties in sequence acquisition and in the integration of statistical regularities over time.

One of the most consistent findings was the presence of generalized response slowing accompanied by reduced implicit learning in the schizophrenia group across both modalities (Figure 1, bottom graph). Participants with schizophrenia exhibited longer reaction times throughout the SRTT, consistent with the well-established psychomotor slowing observed in schizophrenia (Gebreegziabhere et al., 2022). In addition, our findings suggest impairments in probabilistic learning mechanisms, which have also been consistently reported in schizophrenia (Weickert, 2018). Previous studies have shown that individuals with schizophrenia tend to draw conclusions from insufficient evidence, as demonstrated by the beads task, and exhibit deficits in integrating probabilistic contingencies over time (Averbeck et al., 2011). In contrast, the control group demonstrated clear evidence of implicit learning, particularly during motor execution (Figure 1, top graph), with progressively faster reaction times and stable modulation by stimulus type across practice, consistent with the seminal findings of Nissen and Bullemer (1987). More specifically, the Control group showed significant effects of stimulus type under both motor execution and motor imagery, indicating sensitivity to the statistical regularities embedded in the sequence across both modalities. Significant block effects further suggest that performance was modulated over time. Interestingly, under motor imagery, the absence of a stimulus type × block interaction suggests that the relative differences between stimulus types remained stable throughout practice, in contrast to the pattern observed during motor execution. In the schizophrenia group, however, sensitivity to the statistical structure of the sequence was observed only during motor execution, whereas no reliable effects emerged under motor imagery. When reaction times were compared across modalities, this pattern was further reflected by the absence of a main effect of modality and by a less pronounced differentiation between stimulus types, particularly during motor execution.

Furthermore, when reaction times were compared across modalities, the Control group showed a clear modulation of stimulus type as a function of task modality. The distinction between stimulus types was particularly robust during motor execution, whereas under motor imagery these differences were considerably weaker despite the presence of a significant stimulus type × modality interaction. Together with the block-by-block analyses, this pattern suggests that sensitivity to sequence regularities during motor imagery may be less stable over time than during motor execution. Although a significant interaction between stimulus type and modality was also observed in this group, the overall pattern suggests a reduced ability to extract and maintain information about the probabilistic structure of the sequence across both modalities. Similar performance patterns have also been observed in paradigms involving intrinsic variability and motor imagery, as reported in our previous work (Camargo, Passos-Cabral, and Helene, 2026). Together, these findings support the notion that healthy individuals progressively acquire probabilistic regularities through repeated exposure, whereas individuals with schizophrenia exhibit impairments that extend beyond overt motor performance to affect internally generated responses and broader probabilistic learning mechanisms.

A closer examination of performance within the schizophrenia group revealed a distinctive pattern. Although reaction times were modulated by stimulus type, suggesting some degree of sensitivity to task structure, there was no evidence of progressive improvement across blocks. This pattern is consistent with the findings of Weickert et al. (2009), who reported relatively preserved initial acquisition of probabilistic associations in schizophrenia, accompanied by impaired overall performance related to deficits in feedback integration and in the formation of stable stimulus-response associations over time. In this context, performance changes may reflect a general adaptation to the sensorimotor demands of the task rather than the precise encoding of its statistical structure. Under motor imagery, individuals with schizophrenia showed no reliable evidence of learning. Reaction times were not consistently modulated by stimulus type and did not systematically improve across practice blocks. This absence of stable effects suggests greater impairment in internally driven processes involved in mental simulation. These findings are broadly consistent with previous reports describing abnormalities in motor imagery processes in schizophrenia (Maruff et al., 2003).

The correlation analyses further suggest that the behavioral metric derived from the difference between F2 and V2 reaction times may capture distinct aspects of clinical and cognitive functioning depending on task modality. During motor execution, worsening psychopathology, particularly within the PANSS-General domain, was associated with a tendency of progressive reduction in the distinction between stimulus-specific reaction times, indicating diminished sensitivity to probabilistic structure. In contrast, the same cannot be said of the association of PANSS measures and behavioral performance in motor imagery. Regarding cognitive performance, only verbal learning scores from the MCCB showed a tendency of association with the behavioral metric, specifically during motor imagery. This finding suggests that the ability to internally simulate and differentiate sequential regularities may depend, at least partially, on broader cognitive processes related to verbal learning and cognitive integration. Although exploratory in nature, these findings indicate that distinct symptom and cognitive dimensions may differentially relate to probabilistic learning performance across motor execution and motor imagery modalities.

Our findings are broadly consistent with a significant part of Schizophrenia studies which evaluate motor function and cognition, both in human and animal models (Morrens, Hulstijn and Sabbe, 2007; Kalkstein, Hurford and Gur, 2010; Van den Eynde et al., 2014; Chow, Yan and Wu, 2016; Osborne et al., 2020; Goldsmith et al., 2020; Tang et al., 2025). These studies relate psychomotor slowing and cognitive deficits with structural changes and chemical imbalances in the central nervous system. Such alterations have been associated with impairments in reinforcement learning and in the integration of information over time, processes that are directly relevant for the acquisition of probabilistic regularities. The influence of antipsychotic medication on brain connectivity and broader neural organization has also been discussed in the literature (Dodd et al., 2024). In the present study, participants in the schizophrenia group were under stable pharmacological treatment, including both typical and atypical antipsychotics, whereas control participants were screened to exclude neurological and psychiatric conditions and did not use psychotropic medication. Although the present study did not directly investigate neural mechanisms or systematically control for medication effects, the observed behavioral pattern reinforces previous evidence describing implicit learning impairments in schizophrenia (Gebreegziabhere et al., 2022). Importantly, cognitive deficits have also been described in drug-naive first-episode patients as well as in stable-treatment schizophrenia patients (Gebreegziabhere et al., 2022), suggesting that such impairments cannot be fully attributed to medication effects alone. Furthermore, although Pedersen et al. (2008) discussed the possible influence of medication type and dopaminergic blockade on implicit learning, other studies did not identify significant associations between medication use, disease duration, and task performance (Marvel et al., 2007), or between antipsychotic dose and learning outcomes (Weickert et al., 2009). Nevertheless, the absence of a systematic analysis of medication type and chlorpromazine-equivalent dose remains a limitation of the present study. Within this framework, the current findings may reflect a reduced ability to form and maintain stable sequential representations across repeated practice.

From a clinical and translational perspective, the present findings provide further evidence that implicit probabilistic learning paradigms may represent useful tools for investigating cognitive dysfunctions associated with schizophrenia, particularly those involving predictive processing, sequence acquisition, and internally generated responses. The differential patterns observed between motor execution and motor imagery conditions suggest that these paradigms may help characterize distinct cognitive mechanisms underlying the disorder. In particular, the correlation analyses involving PANSS measures yielded promising results, especially the association between PANSS-General symptom severity and reduced sensitivity to probabilistic structure during motor execution. This finding suggests that alterations in implicit probabilistic learning may be meaningfully related to broader clinical manifestations of schizophrenia, potentially reflecting impairments in the integration and adaptive use of environmental regularities over time. In contrast, the MCCB findings were more restricted, with only the verbal learning domain showing a significant association with behavioral performance during motor imagery. Although exploratory, these results suggest that behavioral metrics derived from probabilistic learning paradigms may contribute to the characterization of clinically relevant dimensions of schizophrenia and may eventually support the development of complementary approaches for monitoring disease-related changes or evaluating the effects of cognitive and rehabilitative interventions. Future studies integrating behavioral, electrophysiological, and neuroimaging approaches may further clarify the neural mechanisms underlying these effects and help determine the sensitivity and specificity of these paradigms in clinical contexts.

Several limitations should nevertheless be acknowledged. Differences in response requirements between motor execution and motor imagery conditions may have influenced performance patterns across modalities. In addition, clinical variables such as medication status, symptom heterogeneity, disease duration, and treatment history are inherently difficult to isolate and systematically control in schizophrenia research, and may have contributed to variability in the observed effects. Importantly, the results obtained from the correlation analyses should be interpreted with caution. Although the associations observed with PANSS measures are theoretically meaningful and consistent with the broader behavioral findings, additional studies involving independent cohorts and larger samples will be necessary to determine the robustness, reproducibility, and potential applicability of these effects. The MCCB-related findings should be interpreted even more cautiously, given the more limited pattern of significant associations observed. Future investigations may help establish whether these behavioral markers are sufficiently stable and sensitive to support their use in clinical or translational settings.

## 4. Conclusion

The present study provides evidence that individuals with schizophrenia exhibit impairments in implicit probabilistic sequence learning, characterized by reduced sensitivity to sequence-related regularities and limited adaptation across repeated practice. Importantly, by directly comparing motor execution and motor imagery within the same probabilistic learning paradigm, we demonstrated that these impairments are evident across both modalities. This finding extends previous evidence by revealing a deficit in internally driven learning processes that has received relatively little attention in the schizophrenia literature. Furthermore, the observed associations between behavioral performance and clinical measures, particularly PANSS-General scores, indicate that alterations in implicit probabilistic learning may be meaningfully related to broader symptom dimensions in schizophrenia. Together, these findings reinforce the relevance of probabilistic learning paradigms for investigating cognitive dysfunctions associated with the disorder and support future efforts aimed at characterizing behavioral markers related to predictive processing and adaptive learning mechanisms in schizophrenia.

## Acknowledgement

This work has been developed as part of the activities of FAPESP’s Research, Innovation and Dissemination Center for Neuromathematics (NeuroMat, grant 2013/07699-0) and FAPESP’s São Paulo Excellence Chair for Innovations in Human and Non-Human Animal Communities (grant 2018/18900-1). P.S.d.C. is supported by FAPESP’s grant 2023/17520-9. PRC-P was supported by FAPESP grant 2022/00699-3 and INCT-FAPESP grant 2025/27064-6. J.S.G. is supported by National Coordination of High Education Personnel Formation Programs (CAPES) – Finance Code 001.

## Data availability

The anonymized dataset underlying this study, along with the code used for data processing and analysis, is publicly available at the following link: https://github.com/PauloCabral-hub/Behavioral-Statistical-Learning-in-Schizophreny

## Author Contributions

Y.L.U.: Conceptualization, Methodology, Investigation, Resources, Visualization, Writing—original draft; P.S.d.C.: Conceptualization, Methodology, Investigation, Resources, Visualization, Writing—review & editing; PRC-P: Formal analysis, Visualization, Writing—review & editing; B.R.: Investigation, Visualization, Writing—review & editing; J.S.G.: Investigation, Visualization, Writing—review & editing; A.F.H.: Conceptualization, Methodology, Investigation, Resources, Visualization, Writing—review & editing, Funding acquisition; A.G.: Investigation, Visualization, Project administration; D.A.: Investigation, Supervision, Project administration. All authors have read and agreed to the published version of the manuscript.

