## Supplementary tables for "Impaired Probabilistic Learning deficits in Schizophrenia: A study with Motor Execution and Imagery"

**Reduced Sensitivity to Probabilistic Regularities During Motor Execution and Motor Imagery in Schizophrenia**

**PANSS subscales scores**

**Table S1. PANSS subscales scores for participants in the Schizophrenia Group separated by modality. Mean and standard deviations (SD) are given.**

|  | **Motor Execution**  **(Mean SD )** | **Motor Imagery**  **(Mean SD)** |
| --- | --- | --- |
| PANSS-Positve | 9.8$\pm$2.10 | 11.50$\pm$3.20 |
| PANSS-Negative | 13.20$\pm$3.5 | 13.20$\pm$3.50 |
| PANSS-General | 20.50$\pm$4.80 | 23.20$\pm$5.50 |
| PANSS-Total | 43.5$\pm$7.20 | 50.50$\pm$8.50 |

PANSS - Positive and Negative Symptoms Scale.

**Antipsychotic medications**

**Table S2. Antipsychotic medications used by participants in the Schizophrenia Group. Mean and standard deviations (SD) of chlorpromazine equivalent doses (mg/day) are given.**

| **Antipsychotic Classification** | **Medication** | **Number of patients** | **Chlorpromazine Eq.** |
| --- | --- | --- | --- |
| Typical Antipsychotics | Haloperidol | 6 | 283.4 $\pm$ 165.2 mg |
| Typical Antipsychotics | Haloperidol Decanoate | 4 | 303.0 $\pm$ 176.5 mg |
| Atypical Antipsychotics | Clozapine | 8 | 456.3 $\pm$ 198.7 mg |
| Atypical Antipsychotics | Olanzapine | 4 | 660.4 $\pm$ 225.1 mg |
| Atypical Antipsychotics | Risperidone | 4 | 179.7 $\pm$ 59.9 mg |
| Atypical Antipsychotics | Aripiprazole | 3 | 633.3 $\pm$ 305.5 mg |
| Atypical Antipsychotics | Quetiapine | 3 | 172.9 $\pm$ 140.6 mg |
| Atypical Antipsychotics | Paliperidone | 1 | 13150.0 mg |
| Atypical Antipsychotics | Palmitato de Paliperidona | 1 | 301.2 mg |

Eq. - Equivalent

**MATRICS Consensus Cognitive Battery (MCCB)**

**Table S3: Cognitive performance of participants on the Matricis Consensus Cognitive Battery (MCCB).**

| **id** | **TMT** | **TMT_t** | **BACS** | **BACS_t** | **HVLT** | **HVLT_t** | **DS_for** | **DS_back** | **DS** | **DS_t** | **LNS** | **LNS_t** | **Maze** | **Maze_t** | **BVMT** | **BVMT_t** | **Fluency** | **Fluency_t** | **CPT_2D** | **CPT_3D** | **CPT_4D** | **CPT** | **CPT_t** |
| --- | --- | --- | --- | --- | --- | --- | --- | --- | --- | --- | --- | --- | --- | --- | --- | --- | --- | --- | --- | --- | --- | --- | --- |
| 1 | 40 | 46 | 57 | 54 | 30 | 58 | 10 | 8 | 18 | 55 | 15 | 53 | 19 | 44 | 20 | 37 | 30 | 60 | 4241 | 25676 | 1.1 | 26362 | 53 |
| 2 | 41 | 46 | 40 | 40 | 19 | 29 | 9 | 6 | 15 | 47 | 15 | 53 | 17 | 44 | 25 | 44 | 20 | 41 | 39593 | 18382 | 16893 | 24956 | 50 |
| 3 | 55 | 37 | 35 | 38 | 13 | 15 | 8 | 10 | 18 | 55 | 11 | 45 | 15 | 41 | 13 | 15 | 12 | 27 | 34022 | 13655 | 1 | 2 | 42 |
| 4 | 30 | 54 | 40 | 40 | 22 | 37 | 10 | 9 | 19 | 55 | 10 | 40 | 24 | 50 | 24 | 44 | 21 | 42 | 23941 | 14988 | 1 | 15353 | 39 |
| 10 | 59 | 42 | 52 | 58 | 11 | 23 | 9 | 5 | 14 | 53 | 12 | 50 | 10 | 42 | 11 | 23 | 19 | 46 | 25676 | 18388 | 10676 | 18246 | 48 |
| 11 | 44 | 51 | 30 | 42 | 12 | 23 | 8 | 2 | 10 | 40 | 6 | 37 | 4 | 35 | 6 | 9 | 13 | 36 | 2.0228 | 0.98168 | 0.43121 | 1.452 | 41 |
| 13 | 26 | 60 | 47 | 55 | 19 | 36 | 9 | 8 | 17 | 61 | 13 | 50 | 25 | 60 | 17 | 36 | 18 | 44 | 2.9512 | 2.1782 | 1.2808 | 2.137 | 53 |
| 15 | 44 | 43 | 45 | 45 | 19 | 29 | 8 | 6 | 14 | 47 | 7 | 35 | 12 | 37 | 12 | 15 | 20 | 41 | 2.3403 | 0.87451 | 0 | 1.0716 | 31 |
| 25 | 44 | 43 | 27 | 31 | 17 | 29 | 7 | 5 | 12 | 40 | 10 | 40 | 16 | 41 | 6 | 1 | 24 | 49 | 2.3734 | 1.7129 | 0.53896 | 1.5417 | 39 |
| 26 | 34 | 57 | 34 | 44 | 20 | 43 | 9 | 10 | 19 | 61 | 17 | 63 | 22 | 56 | 23 | 49 | 26 | 57 | 3.6193 | 2.4604 | 0.79763 | 2.2924 | 53 |
| 27 | 20 | 61 | 54 | 51 | 28 | 51 | 10 | 8 | 18 | 55 | 17 | 60 | 24 | 50 | 16 | 22 | 25 | 50 | 3.4022 | 2.7805 | 1.5814 | 2.588 | 53 |
| 28 | 52 | 47 | 28 | 39 | 26 | 56 | 6 | 5 | 11 | 45 | 13 | 50 | 4 | 35 | 25 | 49 | 18 | 44 | 1.4989 | 2.1205 | 1.4995 | 1.7063 | 48 |
| 35 | 48 | 41 | 34 | 36 | 16 | 22 | 5 | 4 | 9 | 36 | 11 | 45 | 11 | 36 | 12 | 15 | 17 | 36 | 1.9321 | 1.1456 | 0.2044 | 1.094 | 31 |
| 36 | 30 | 54 | 34 | 36 | 15 | 22 | 7 | 7 | 14 | 47 | 10 | 40 | 13 | 37 | 9 | 10 | 28 | 56 | 3.6193 | 2.6803 | 0.68117 | 2.3269 | 50 |
| 37 | 86 | 13 | 31 | 34 | 6 | 1 | 4 | 7 | 11 | 40 | 3 | 24 | 15 | 41 | 12 | 15 | 14 | 30 | 1.3503 | 0.98168 | 1.7562 | 1.3627 | 36 |
| 12 | 35 | 50 | 38 | 40 | 21 | 37 | 9 | 9 | 18 | 55 | 13 | 45 | 12 | 37 | 13 | 15 | 25 | 50 | 3.9593 | 3.0865 | 1.0681 | 2.7046 | 56 |
| 32 | 77 | 19 | 26 | 31 | 23 | 44 | 10 | 7 | 17 | 55 | 8 | 35 | 6 | 30 | 25 | 44 | 10 | 23 | 1.781 | 2.464 | 1.199 | 1.814 | 42 |
| 20 | 92 | 9 | 25 | 29 | 16 | 22 | 4 | 2 | 6 | 26 | 6 | 30 | 4 | 28 | 9 | 10 | 7 | 21 | 1.667 | 1.921 | 0.19 | 1.259 | 34 |
| 21 | 57 | 75 | 55 | 51 | 21 | 37 | 10 | 9 | 19 | 55 | 17 | 60 | 26 | 53 | 15 | 22 | 26 | 52 | 3.959 | 1.279 | 1.841 | 2.359 | 50 |
| 22 | 58 | 34 | 32 | 36 | 19 | 29 | 5 | 5 | 10 | 36 | 9 | 40 | 6 | 30 | 10 | 10 | 15 | 32 | 2.092 | 1.094 | 0.773 | 1.319 | 36 |
| 16 | 34 | 52 | 36 | 38 | 19 | 29 | 8 | 4 | 12 | 40 | 9 | 40 | 10 | 34 | 10 | 10 | 16 | 34 | 3.959 | 2.247 | 1.195 | 2.467 | 50 |
| 17 | 37 | 50 | 38 | 40 | 19 | 29 | 9 | 5 | 14 | 47 | 4 | 24 | 23 | 50 | 9 | 10 | 20 | 41 | 0.955 | 1.365 | 0.421 | 0.913 | 31 |
| 8 | 59 | 32 | 46 | 45 | 23 | 44 | 8 | 6 | 14 | 47 | 10 | 40 | 14 | 39 | 13 | 15 | 15 | 32 | 4.241 | 2.611 | 1.28 | 2.71 | 56 |
| 19 | 70 |  | 29 |  | 20 |  | 6 | 4 | 10 |  | 15 |  | 15 |  | 16 |  | 17 |  | 4.241 | 1.1456 | 1.4517 | 2.279 |  |
| 6 | 27 | 57 | 47 | 45 | 20 | 29 | 9 | 6 | 15 | 47 | 13 | 45 | 23 | 48 | 28 | 51 | 18 | 38 | 3.9593 | 3.6777 | 1.16 | 2.5457 | 50 |
| 18 | 31 | 54 | 44 | 45 | 19 | 29 | 8 | 7 | 15 | 47 | 13 | 45 | 25 | 53 | absent | absent | 23 | 47 | 3.62 | 2.37 | 1.53 | 2.51 | 53 |
| 34 | 27 | 57 | 32 | 36 | 21 | 37 | 7 | 8 | 15 | 47 | 8 | 35 | 21 | 47 | absent | absent | 21 | 43 | 3.1206 | 1.4501 | 0.41025 | 2.891 | 56 |
| 9 | 53 | 47 | 30 | 34 | 25 | 44 | absent | absent | 16 | 47 | 11 | 40 | 12 | 37 | 14 | 22 | 18 | 38 | 2.3734 | 1.1545 | 1.1991 | 1.575 | 36 |
| 14 | 19 | 63 | 55 | 51 | 31 | 58 | 6 | 6 | 12 | 40 | 14 | 53 | 23 | 50 | 35 | 72 | 22 | 45 | 4.24 | 2.08 | 1.46 | 2.6 | 53 |
| 7 | 40 | 48 | 32 | 36 | 23 | 44 | 8 | 2 | 10 | 36 | 9 | 40 | 12 | 37 | absent | absent | 18 | 38 | 2.55 | 2.12 | 0.6 | 1.76 | 42 |
| 30 | 19 | 63 | 46 | 45 | 27 | 51 | 8 | 10 | 18 | 55 | 11 | 45 | 24 | 50 | 30 | 58 | 21 | 43 | 2.46 | 2.36 | 1.84 | 2.22 | 48 |

id = patient; TMT = Trail Making Test; BACS = Brief Assessment of Cognition in Schizophrenia (Symbol Coding); HVLT = Hopkins Verbal Learning Test–Revised; DS Forward = Digit Span Forward; DS Backward = Digit Span Backward; DS = Total Digit Span score; LNS = Letter-Number Sequencing; Maze = Neuropsychological Assessment Battery Mazes Test; BVMT-R = Brief Visuospatial Memory Test–Revised; Verbal Fluency = Category Fluency (Animals); CPT 2D/3D/4D = Continuous Performance Test–Identical Pairs (2-, 3-, and 4-digit conditions); CPT = CPT-IP composite score; t = standardized T score (mean = 50, standard deviation = 10). Missing values are indicated by “—”. “absentl” indicates that the instrument was not administered to the participant.

**Statistical Analysis - Tables**

**Table S4: Assessment of normality assumptions for reaction time data used in the two-way repeated-**

**measures ANOVA of Figure 3.**

| **stim_type** | **W** | **pval** | **normal** | **set** | **group** |
| --- | --- | --- | --- | --- | --- |
| f1 | 0.872386 | 1,49E+01 | FALSE | schizophrenia | 1 |
| f2 | 0.722507 | 2,47E-03 | FALSE | schizophrenia | 1 |
| v2 | 0.933121 | 2,70E+03 | FALSE | schizophrenia | 1 |
| v3 | 0.893358 | 7,60E+01 | FALSE | schizophrenia | 1 |
| f1 | 0.956891 | 2,37E+04 | FALSE | schizophrenia | 2 |
| f2 | 0.981902 | 4,59E+05 | TRUE | schizophrenia | 2 |
| v2 | 0.947293 | 7,79E+03 | FALSE | schizophrenia | 2 |
| v3 | 0.982617 | 4,93E+05 | TRUE | schizophrenia | 2 |
| f1 | 0.891238 | 5,60E-01 | FALSE | control | 1 |
| f2 | 0.929050 | 4,40E+01 | FALSE | control | 1 |
| v2 | 0.892839 | 6,61E-01 | FALSE | control | 1 |
| v3 | 0.794881 | 1,73E-04 | FALSE | control | 1 |
| f1 | 0.637659 | 2,37E-08 | FALSE | control | 2 |
| f2 | 0.605697 | 5,53E-09 | FALSE | control | 2 |
| v2 | 0.711450 | 1,01E-06 | FALSE | control | 2 |
| v3 | 0.627336 | 1,47E-08 | FALSE | control | 2 |

Stim_type = event type; f1 = event F1; f2 = event F2; v12 = event v2; v3 = event v3; W = Shapiro-Wilk test statistic;

*pval* = *p* value; set = experimental groups.

**Table S5: Assessment of normality assumptions for reaction time data used in the two-way repeated-**

**measures ANOVA after rank-transformation of Figure 3.**

| **stim_type** | **W** | **pval** | **normal** | **set** | **group** |
| --- | --- | --- | --- | --- | --- |
| f1 | 0.955228 | 0.027612 | FALSE | schizophrenia | 1 |
| f2 | 0.955228 | 0.027612 | FALSE | schizophrenia | 1 |
| v2 | 0.955228 | 0.027612 | FALSE | schizophrenia | 1 |
| v3 | 0.955228 | 0.027612 | FALSE | schizophrenia | 1 |
| f1 | 0.955107 | 0.019175 | FALSE | schizophrenia | 2 |
| f2 | 0.955107 | 0.019175 | FALSE | schizophrenia | 2 |
| v2 | 0.955107 | 0.019175 | FALSE | schizophrenia | 2 |
| v3 | 0.955107 | 0.019175 | FALSE | schizophrenia | 2 |
| f1 | 0.954725 | 0.001722 | FALSE | control | 1 |
| f2 | 0.954725 | 0.001722 | FALSE | control | 1 |
| v2 | 0.954725 | 0.001722 | FALSE | control | 1 |
| v3 | 0.954725 | 0.001722 | FALSE | control | 1 |
| f1 | 0.954725 | 0.001722 | FALSE | control | 2 |
| f2 | 0.954725 | 0.001722 | FALSE | control | 2 |
| v2 | 0.954725 | 0.001722 | FALSE | control | 2 |
| v3 | 0.954725 | 0.001722 | FALSE | control | 2 |

Stim_type = event type; f1 = event F1; f2 = event F2; v12 = event v2; v3 = event v3; W = Shapiro-Wilk test statistic;

*pval* = *p* value; set = experimental groups.

**Table S6: Results of two-way repeated-measures ANOVA based on permutation test for reaction times across blocks and stimulus types in the Schizophrenia and Control groups (Figure 3).**

| **Source** | **SS** | **ddof1** | **ddof2** | **MS** | **F** | **p-unc** | **p-GG-corr** | **ng2** | **eps** | **data_set** | **group** |
| --- | --- | --- | --- | --- | --- | --- | --- | --- | --- | --- | --- |
| block | 0.105734 | 4 | 44 | 0.026434 | 2.429.553 | 0.0551 | 0.122582 | 0.048630 | 0.418171 | schizophreny | 1 |
| stim_type | 0.037045 | 3 | 33 | 0.012348 | 1.521.829 | 0.2193 | 0.236677 | 0.017594 | 0.770302 | schizophreny | 1 |
| block * stim_type | 0.023057 | 12 | 132 | 0.001921 | 0.511369 | 0.9126 | 0.692168 | 0.011024 | 0.272238 | schizophreny | 1 |
| block | 0.018763 | 4 | 48 | 0.004691 | 0.386807 | 0.8224 | 0.727370 | 0.010908 | 0.624138 | schizophreny | 2 |
| stim_type | 0.028321 | 3 | 36 | 0.009440 | 1.308.529 | 0.2870 | 0.288974 | 0.016373 | 0.754642 | schizophreny | 2 |
| block * stim_type | 0.037719 | 12 | 144 | 0.003143 | 0.949044 | 0.5030 | 0.422736 | 0.021688 | 0.232385 | schizophreny | 2 |
| block | 0.050906 | 4 | 76 | 0.012726 | 1.981.761 | 0.1032 | 0.130150 | 0.016353 | 0.715610 | control | 1 |
| stim_type | 0.206663 | 3 | 57 | 0.068888 | 5.481.812 | 0.0019 | 0.008926 | 0.063225 | 0.641813 | control | 1 |
| block * stim_type | 0.070184 | 12 | 228 | 0.005849 | 1.212.247 | 0.2675 | 0.312293 | 0.022407 | 0.344514 | control | 1 |
| block | 2.461.870 | 4 | 76 | 0.615467 | 5.464.565 | 0.0001 | 0.015765 | 0.031763 | 0.375234 | control | 2 |
| stim_type | 0.091186 | 3 | 57 | 0.030395 | 1.721.057 | 0.1621 | 0.189462 | 0.001214 | 0.720125 | control | 2 |
| block * stim_type | 0.080192 | 12 | 228 | 0.006683 | 0.850771 | 0.6144 | 0.405279 | 0.001067 | 0.121670 | control | 2 |

Separate analyses were conducted for each group (Schizophrenia and Control) and experimental set (1 and 2). The within-subject factors were Block and Stimulus Type. SS = sum of squares; ddof1 = numerator degrees of freedom; ddof2 = denominator degrees of freedom; MS = mean square; F = F statistic; p-unc = uncorrected p-value; p-GG-corr = Greenhouse–Geisser corrected p-value; ng² = generalized eta squared effect size; eps = Greenhouse–Geisser epsilon. Stimulus types included f1 (event F1), f2 (event F2), v2 (event V2), and v3 (event V3). Significant effects after Greenhouse–Geisser correction are indicated by p-GG-corr < .05.

**Table S7: Assessment of normality assumptions for reaction time data used in the two-way repeated-**

**measures ANOVA after rank-transformation of Figure 4.**

| **stim_type** | **W** | **pval** | **normal** | **data_set** | **group** |
| --- | --- | --- | --- | --- | --- |
| f1 | 0.936458 | 0.453598 | TRUE | s | g |
| f2 | 0.918305 | 0.272185 | TRUE | s | g |
| v2 | 0.943192 | 0.540490 | TRUE | s | g |
| v3 | 0.849262 | 0.035951 | FALSE | s | g |
| f1 | 0.951245 | 0.617263 | TRUE | s | g |
| f2 | 0.962801 | 0.796235 | TRUE | s | g |
| v2 | 0.915602 | 0.218687 | TRUE | s | g |
| v3 | 0.920914 | 0.258073 | TRUE | s | g |
| f1 | 0.967072 | 0.692237 | TRUE | s | g |
| f2 | 0.885690 | 0.022427 | FALSE | s | g |
| v2 | 0.966327 | 0.676216 | TRUE | s | g |
| v3 | 0.953658 | 0.426043 | TRUE | s | g |
| f1 | 0.895600 | 0.034131 | FALSE | s | g |
| f2 | 0.906467 | 0.054604 | TRUE | s | g |
| v2 | 0.874259 | 0.013971 | FALSE | s | g |
| v3 | 0.900672 | 0.042453 | FALSE | s | g |

Stim_type = event type; f1 = event F1; f2 = event F2; v12 = event v2; v3 = event v3; W = Shapiro-Wilk test statistic;

*pval* = *p* value; Data_set and group identify the corresponding experimental dataset and participant group.
